# Social network heterogeneity is essential for contact tracing

**DOI:** 10.1101/2020.06.05.20123141

**Authors:** Bjarke Frost Nielsen, Kim Sneppen, Lone Simonsen, Joachim Mathiesen

**Affiliations:** Niels Bohr Institute, University of Copenhagen, 2100 Copenhagen, Denmark; Department of Science and Environment, Roskilde University, 4000 Roskilde, Denmark

**Keywords:** COVID-19, Epidemiological modeling, Social structure, Contact tracing

## Abstract

Contact tracing is suggested as an effective strategy for controlling an epidemic without severely limiting personal mobility. Here, we explore how social structure affects contact tracing of COVID-19. Using smartphone proximity data, we simulate the spread of COVID-19 and find that heterogeneity in the social network and activity levels of individuals decreases the severity of an epidemic and improves the effectiveness of contact tracing. As a mitigation strategy, contact tracing depends strongly on social structure and can be remarkably effective, even if only frequent contacts are traced. In perspective, this highlights the necessity of incorporating social heterogeneity into models of mitigation strategies.

Significance Statement
The COVID-19 epidemic has put severe limitations on individual mobility in the form of lockdowns and closed national borders. Mitigation strategies permitting individual mobility while limiting disease spreading are needed, and contact tracing is a potentially effective example of such a strategy. We use smartphone proximity data to monitor contacts between people, and find that contact tracing is highly dependent on social structure, being very effective on real contact networks. This shows that mitigation of COVID-19 may be possible with contact tracing, and that epidemiological models must incorporate social network structure.

---

**E**pidemics are typically modeled by well-mixed compartmental models (1–4) where some degree of social heterogeneity can be introduced by sub-dividing the model population according to e.g. age, occupation, household and social spheres (5–10). Despite the possibility of adjusting interaction rates between sub-populations, well-mixed models may fail to predict the evolution of an epidemic when social interactions are spatio-temporally restricted (10), like in real *contact networks*. The social interactions of individuals tend to follow a characteristic pattern; you meet the same people at specific times during a week, form groups based on social preferences and, in addition, the number of weekly contacts varies significantly from person to person – phenomena which contribute to transmission heterogeneity. In a well-mixed model – even if stratified by e.g. age – your contacts are essentially drawn at random at each new instant, neglecting the monotony seen in real networks. In previous epidemics, perhaps most notably the Ebola epidemic of 2014-16, transmission heterogeneity and mobility patterns have been shown to be decisive factors (11). Lately, contact tracing – a mitigation strategy which relies directly on the contact network structure – has been the center of much attention due to its promises of epidemic control in a relatively open society (12–17). In order to asses contact tracing strategies, detailed information on contact networks is indispensable, and the usual well-mixed approach is inadequate – even more so than when modelling unmitigated spreading (18, 19).

In this paper, we utilize Bluetooth proximity data obtained from a cohort of university students at a large European university (see *Methods* for details). These data are similar in nature to the sort of readings one might obtain from contact tracing smartphone applications (20), meaning that they provide a useful virtual laboratory for contact tracing. Whereas our data only comprise a section of the total contact network of each participant, they display well-defined and robust heterogeneity, the effects of which can be studied, and compared with analogous homogenized networks.

The participant group is homogeneous in age and occupation, and would consequently be treated as undifferentiated in typical epidemiological models – an assumption that we can directly probe the validity of, in the context of contact tracing. Specifically, we will consider the effects of **contact heterogeneity** on the spread of COVID-19. For that purpose, we introduce three degrees of heterogeneity: **i)** an unaltered real network. **ii)** an edge-swapped version of the network (21), retaining contact heterogeneity but eliminating group formation preferences, including spatial preferences. **iii)** a randomized network, retaining only the overall (mean) contact frequency, but eliminating heterogeneity. This allows us to investigate questions such as whether it affects the spread of COVID-19 if some people are more social than others or if the the social network has distinct sub-communities. Furthermore, we suggest a realistic and easy-to-implement **contact tracing** scheme. We quantify the influence of contact heterogeneity *on* contact tracing in terms of two key parameters, the frequency by which symptomatic individuals are tested and the duration of social contacts with exposed individuals that would trigger a self-quarantine. The latter is especially interesting, since it is a directly controllable parameter when e.g. designing contact tracing smartphone applications (20).

## Materials and Methods

We analyze social proximity and contact dynamics from data collected by smartphones distributed among around 1000 participants (undergraduate students at the Technical University of Denmark (22, 23)). The smartphones were equipped with an application that collected communication in the form of call and text messaging logs, geolocation (GPS coordinates) and social proximity using the Bluetooth port. All smartphones in the study were programmed to open their Bluetooth ports every 5 minutes to scan for nearby devices included in the study and to record the GPS coordinates. The data we consider have been collected over a period of two years, 2013-2015.

The distance between participants is inferred from the strength (RSSI) of the Bluetooth signal being sent between the devices. The signal strength can resolve distances in the range of *≤* 1 meter to approximately 10-15 meters (24). We define a *contact* between two individuals whenever the Bluetooth signal strength between their respective devices exceeds *−*85dBm. This definition of contact captures essentially all *≤* 1m interactions while excluding a large portion of the 3m interactions and above (24), in line with the recommendations of public health authorities (25, 26). This allows us to create a well-defined time-dependent contact network where individuals are represented by nodes and social contact by time-dependent links, similarly to the work of (27). The link activity, or the contact between nodes, is resolved in temporal windows of 5 minutes. This time-dependent contact network will be the basis for our modeling of the transmission of COVID-19.

We model the spread of COVID-19 by an agent-based model (where the study participants serve as the agents) with five states: **S**usceptible to the disease, **E**xposed, **P**re-symptomatic (but infectious), **I**nfected (potentially with symptoms) and **R**ecovered/Removed. In the absence of contact tracing (described below), the P and I states are identical, in that an individual in one of these states can infect others. Aside from these mutually exclusive states, persons can also be flagged as **Q**uarantined. When a susceptible person comes into contact with a person in the **I** or **P** state, there is a probability *p*_inf_ of transmission of the disease in each 5-minute window. The basic model (without contact tracing) thus has four parameters: Transmission probability upon contact *p*_inf_, a time-scale characterizing the exposed state *τ*_E_, a time-scale characterizing the pre-symptomatic state *τ*_P_ and a time-scale characterizing the infected state *τ*_I_.

As shown in the model illustration of Fig. 2, the incubation time is assumed to be gamma-distributed with a mean of 3.6 days, of which 1.2 days comprise the presymptomatic infectious state. The infectious state, where symptoms may be displayed, is set to be 5 days. The last parameter in the model, the transmission probability in each window of time, is fitted to reproduce a daily growth rate. of 23% in the early epidemic, based on estimates from (28).

**Fig 1.**
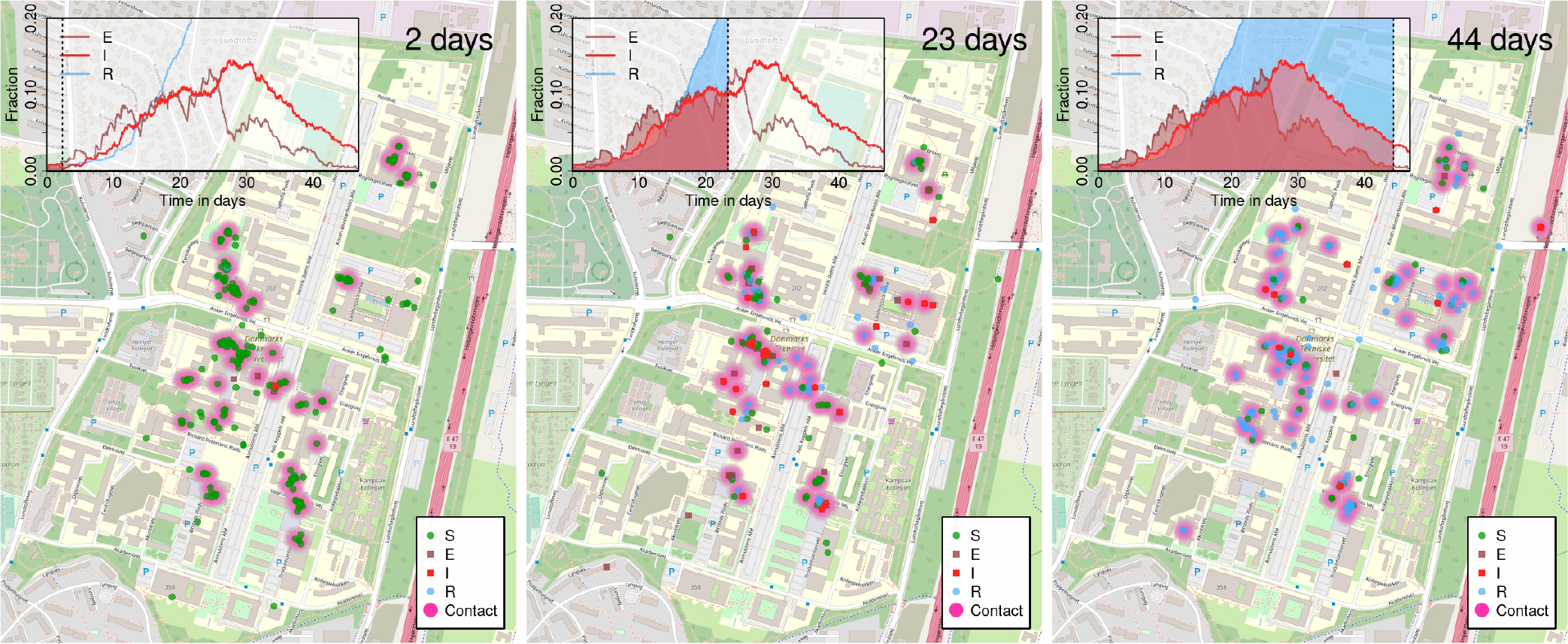
Simulating the spread of COVID-19 on the contact network. Here, a zoom on the geographical positions of individuals (based on GPS coordinates) during a typical work day and for a representative run of the epidemic model. Regions of contact (defined by signal strength exceeding the *−*85dBm cutoff) are shown as diffuse clouds of pink. Snapshots shown are at day 2, 23 and 44 of the outbreak.

**Fig 2.**
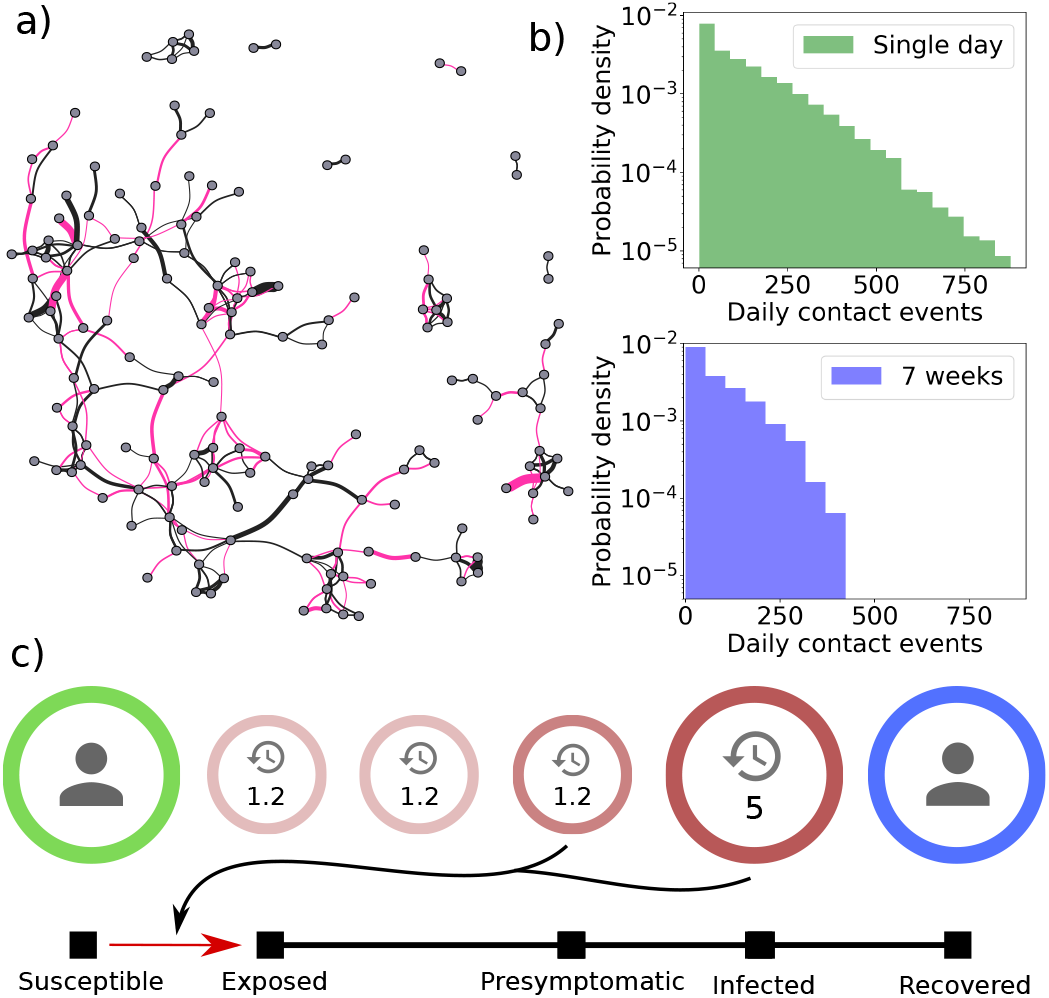
**a)** A small subset of a contact network for one week. Link thickness indicates the cumulative contact time, with links with less than 2 hours cumulative activity being omitted. Black lines represent the links recurring from the previous week, whereas the red lines are new links. **b)** Top: Histogram of contact events over a single day (semi-logarithmic plot). The coefficient of variation is *c*_*V*_ = 1.03 and the mean is *µ* = 131. Bottom: Histogram of contact events over a seven week period, divided by the number of days to obtain an average daily rate (semi-logarithmic plot). Here *c*_*V*_ = 0.95 and *µ* = 86. Both plots show a marked heterogeneity, demonstrating that contact heterogeneity is approximately a quenched disorder on the timescale of a few weeks. **c)** Our agent-based model of COVID-19 spreading on a contact networkIndividuals in the **S**usceptible state may be exposed by those in the **P**resymptomatic as well as **I**nfected states. The Exposed-Presymptomatic triplet of states together comprise the gamma-distributed incubation period.

By employing two different ways of *shuffling* the network con nections (edges), we study the effects of contact heterogeneity on the one hand, and spatial constraints and group formation on the other. The first method, *edge swapping*, preserves the degree of connectivity of each person (node), while destroying any spatial and group formation preference (21). The second method, *randomization*, preserves only the overall connectivity level in each window of time, but homogenizes the number of contacts for each person. In Fig. 3, we plot an epidemic trajectory averaged over 20 simulations.

**Fig 3.**
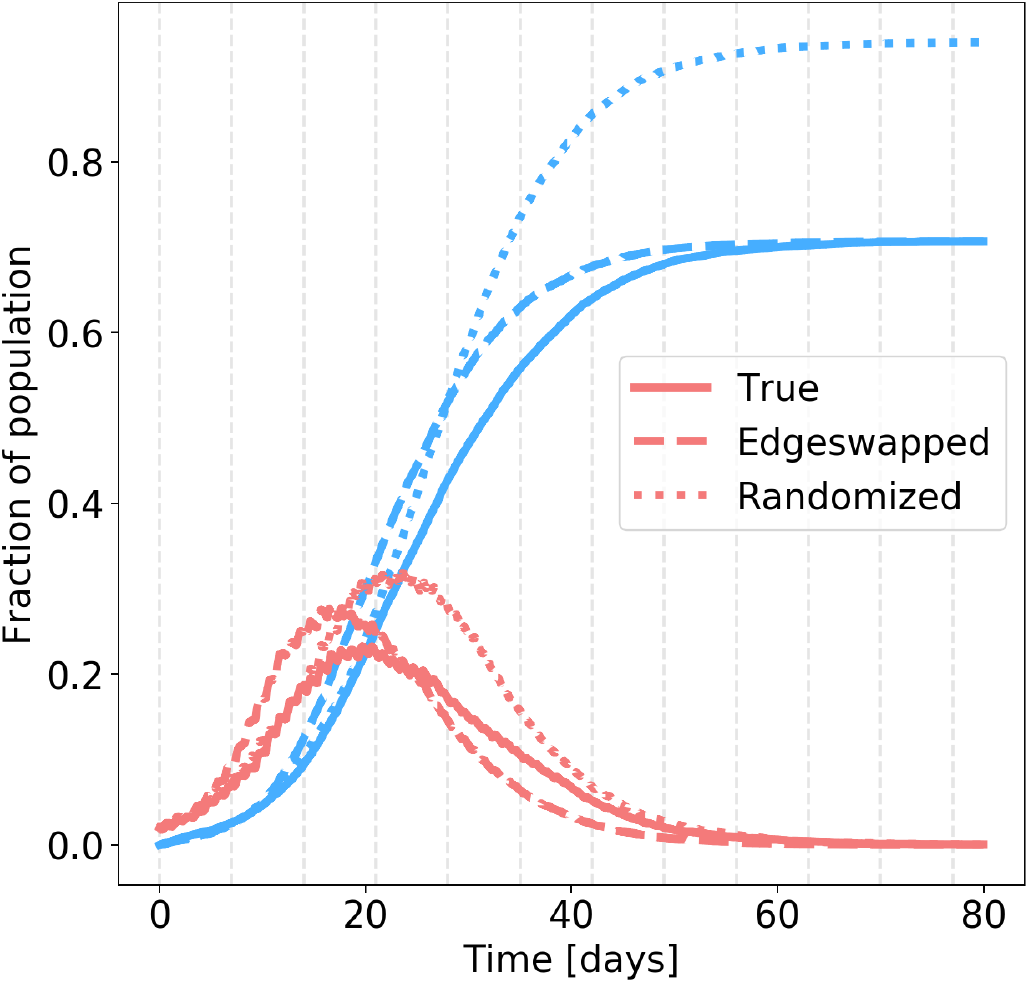
Comparison of exposed + presymptomatic + infected (**red**) and recovered (**blue**) individuals in the three networks types. Each curve represents an ensemble average over 20 simulations. *True network* (full lines): Total fraction exposed: 71%. Infection peak: 28%. *Edgeswapped network* (dashed lines): Total fraction exposed: 71%. Infection peak: 33%. c) *Randomized network* (dotted lines): Total fraction exposed: 94%. Infection peak: 37%. The infection probability per 5-minute contact is *p*_inf_ = 0.5%, fitted to produce a daily growth rate of 23% in the early epidemic, for the true network.

### Contact tracing

The entire scheme around contact tracing consists of two parts: *regular testing* of symptomatic individuals (with a testing probability *p*test *<* 1) and the contact tracing algorithm itself, which is activated once an individual tests positive. Once a positive individual is found by *regular testing*, their recent contacts are put in quarantine for a specified time (5 days as suggested by (17)) and tested once the quarantine period has elapsed (before potential release). In other words, the contact tracing scheme proceeds as follows:

- For each individual, a list of contact events is kept. When a person is tested positive, all contacts older than 5 days (the *retention time*) are discarded, the person is quarantined for 5 days, and all individuals on the contact list are traced.
- If a traced individual has been in contact with the positive person for longer than a certain *contact threshold*, the traced individual is also quarantined for 5 days.
- After the quarantine period has elapsed, the individual is tested. If negative, the individual is released. Otherwise a new 5-day quarantine is issued.

The quarantine is assumed to be instantaneous and a quarantined person has no contact with others. The two simplest assumptions regarding the *regular testing* scheme are that symptomatic individuals are tested at a constant *rate* (throughout their illness), *or* that they have a *fixed probability* of being tested when first developing symptoms. We have compared the two approaches and found that they perform comparably (see Supporting Information), and thus we work with the fixed probability scheme here, for simplicity.

## Results

### Heterogeneous activity levels

The distribution of the number of daily contact events for all individuals in our study is shown in Fig. 2. The distribution reflects a marked heterogeneity in activity levels, characterized by an exponential shape (see Fig. 2b) with a coefficient of variation of 1.03 and a mean of 131. Even more importantly, a significant degree of contact heterogeneity is retained, albeit with some attenuation, when exploring an entire 7-week window. Here the coefficient of variation is 0.95, still close to the value for an exponential distribution, and the mean is 86. It is clear that extreme social behaviour becomes less frequent over this longer time-window, reflecting that individuals do not come into university every single day, with the mean value of 86 corresponding to being inactive on 34% of workdays. However, the significant degree of contact heterogeneity still present shows that it approximately represents a *quenched disorder*, which affects the entire epidemic trajectory and does not simply average out over the course of an epidemic. In the following we explore the profound consequences of contact heterogeneity.

### Heterogeneity reduces epidemic severity

In Fig. 3, we show the simulated evolution of COVID-19 on three different contact networks: The true network (unshuffled), the edge-swapped and the fully randomized network where each person is assigned an average contact frequency. Each trajectory is averaged over 20 runs, each similar in nature to the one shown in the inserts of Fig. 1.

The overall findings are that:

- The total number of exposed individuals is very sensitive to heterogeneity in activity levels, but *not* to network structure. Heterogeneous activity evidently *prevent* the disease from spreading to all parts of the network, with the total fraction exposed reaching 71% in the true network and 94% in the randomized network.
- The infection peak, on the other hand, is sensitive to spatial effects *as well as* contact heterogeneity. As such, the peak load increases by some 5 percentage points when
- spatial structure is destroyed, and by 9 percentage points when contact structure is homogenized as well.
- Overall, the group formation and spatial structure only has the effect of slowing the progression somewhat, but does not affect the attack rate.

### Repeated contacts are essential for contact tracing

The effects of contact heterogeneity and social preference on contact tracing can be probed by utilizing the same two modes of network re-shuffling as in the previous section. In Fig. 4 we compare how the contact tracing algorithm performs on the three networks. It is clear that the epidemic trajectory of the *true* network is drastically altered by contact tracing, with the attack rate and infection peak being profoundly attenuated. In the two shuffled networks, on the other hand, we see very little benefit from contact tracing. This leads to the conclusion, that contact tracing depends strongly on social network structure. Both edge-swapping and randomization reduce contact monotonicity. To quantify this, we find that the median fraction of contacts (of at least 60 minutes cumulative duration) which are repeated from one week to the next is 30% in the real network, while edge-swapping reduces this number to zero. Evidently, this *repetition* of contacts is necessary for contact tracing.

**Fig 4.**
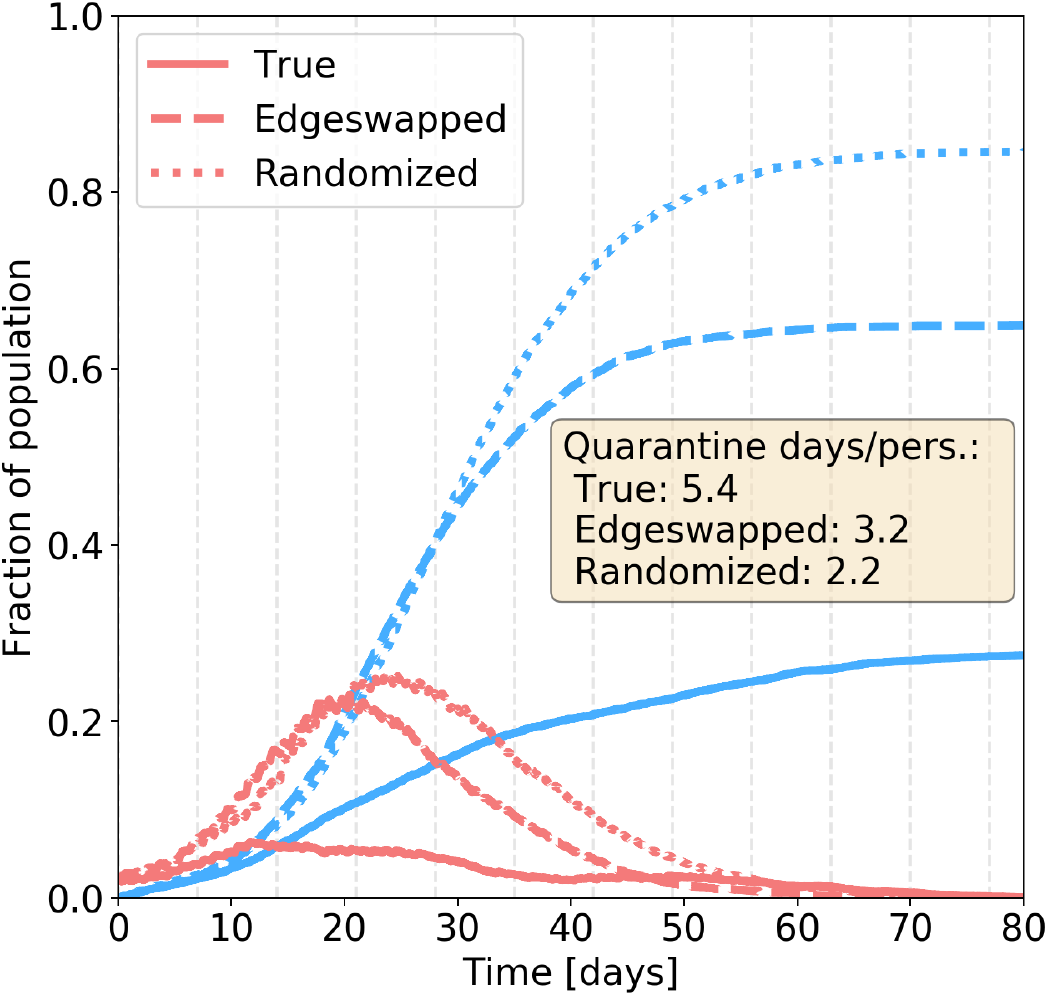
Contact tracing. Comparison of exposed + presymptomatic + infected (**red**) and recovered (**blue**) individuals in the three networks types. Disease parameters are identical to those of Fig. 3. The contact threshold for quarantining is approximately 2 hours (125 minutes) while the testing probability is set at 25%.

### Estimating an optimal contact threshold for efficient trac ing

When making public health decisions about COVID-19 mitigation schemes, it is first and foremost important to have reliable predictions regarding the effectivity of the schemes. Next, however, it is advantageous to identify the parameters which influence the effectiveness. Some of these parameters are beyond our control – for example properties which are intrinsic to SARS-CoV-2 – while others can be partially controlled, or even constitute design decisions on our part.

In the two following sections, we explore two central parameters, namely the *testing probability* and *contact threshold*. The former determines the probability of being tested if sick, while the latter determines how readily quarantines are issued when contact with an infectious person has been established.

### The testing probability

The regular testing required in contact tracing is determined by a *testing probability* which reflects several factors not individually modeled here, such as general testing availability, symptom development and willingness to participate in testing. In Fig. 5a, we explore the influence of the testing probability on the peak load in terms of quarantined and exposed individuals.

**Fig 5.**
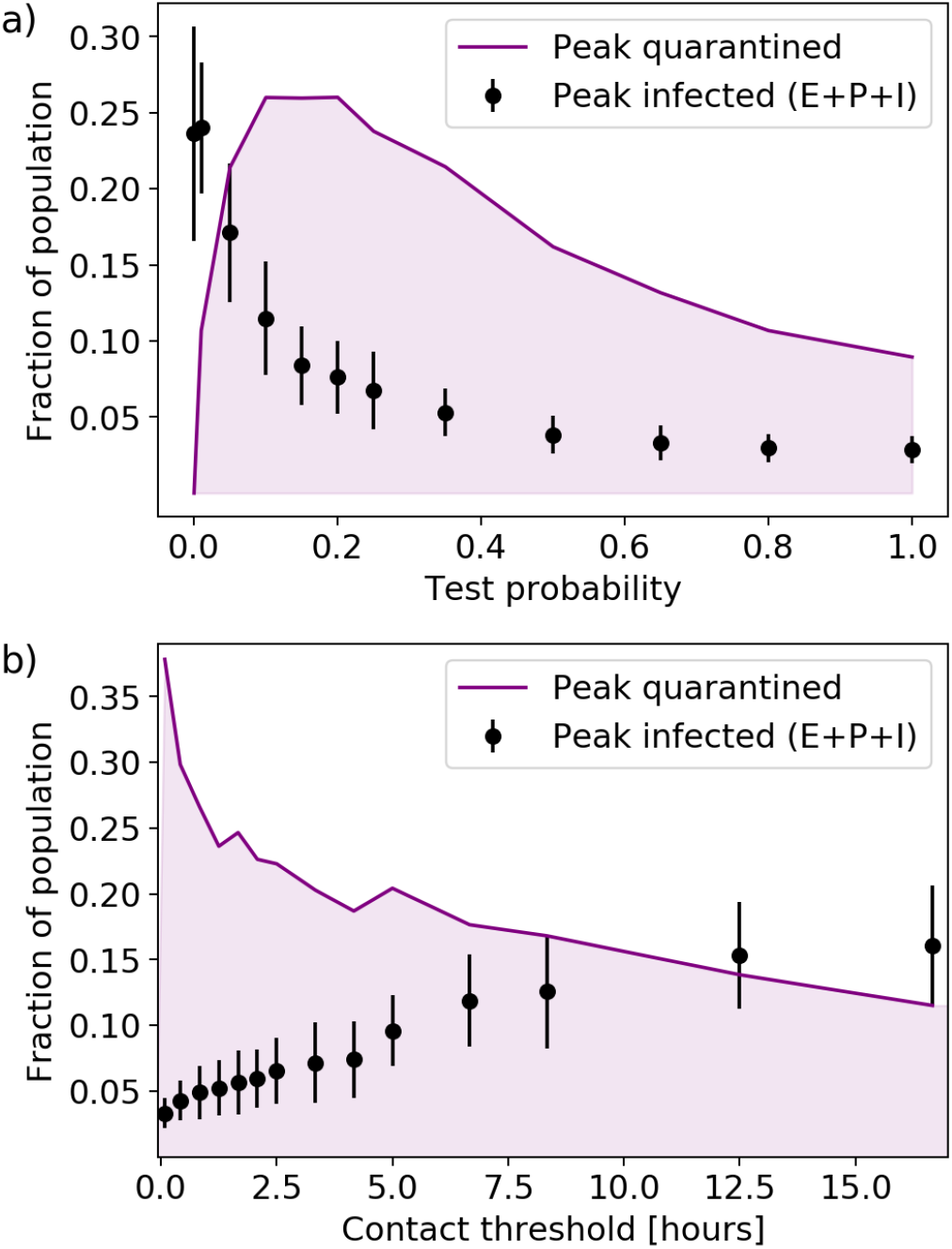
Contact tracing effectiveness. Disease parameters are identical to those of Fig. 3. a) Testing probability vs peak infection and quarantine fraction. The contact threshold is set at 125 minutes. b) Contact threshold vs peak infection and quarantine fraction. The testing probability is set at 25%. For each value of the parameter, 50 simulations were run. The error bars indicate the standard deviation of outcomes of individual simulations.

Unsurprisingly, the quarantine fraction vanishes at very low testing probabilities, where the infection peak attains its maximal value. While the infection load is a decreasing function of testing, the quarantine fraction does not display a simple monotonic response to an increase in testing. Rather, it attains a maximum around 10%, followed by a gradual decline. This clearly shows that changes in testing availability should go hand-in-hand with considerations of the influence on the quarantine fraction, and that the relation is nontrivial.

### The contact threshold

When performing contact tracing, it is necessary to define a *contact threshold*, meaning the minimum duration of proximity between an infectious and a susceptible person which results in quarantine. Setting a low contact threshold will thus intuitively lead to a large fraction of contact persons being placed in quarantine, when a positive individual is found. In Fig. 5b, the contact threshold is shown to have a profound effect on the infection peak as well as the peak fraction of the population in quarantine. As intuition would have it, the infection peak is clearly an increasing function of the contact threshold, while the quarantine fraction decreases. Above a contact threshold of approximately two hours of cumulative proximity, the quarantine fraction decreases only slowly. The peak infection load, on the other hand, increases steadily with the threshold, with a reduction of the epidemic peak down from 25% in the absence contact tracing to 8% when requiring at least 4 hours of cumulative contact within a 5 day window before quarantining. This goes to show that contact tracing is effective, *even if* it is only possible to locate frequent contacts (and 25% of the infected people).

## Discussion

This paper explores the effect of contact heterogeneity on the dynamics and mitigation of an epidemic. In general, we observe that the social activity level of individuals is well approximated by an exponential distribution (see Fig. 2). This observation is consistent with the findings of (5), where a coefficient of variation of social contacts of about 0.8 was reported for people between 20 and 30 years. Further, person-specific social activity remains constant over longer time intervals where both the 1 day and the 7 week activity patterns have coefficients of variations close to 1. Thus the social activity represents a quenched disorder that significantly impedes the spread of the disease and makes the mitigation by contact tracing more efficient. In comparison with our results, traditional well-mixed S(E)IR models overestimate the severity of the epidemic, or, conversely, lead to an underestimation of transmission risk when fitted to an epidemic trajectory. In a previous modelling study (29), it was shown that heterogeneity in the *susceptibility* of individuals likewise reduces the overall severity.

Recently, several studies have found significant heterogeneity in COVID-19 transmission (30, 31). Relatedly, it was shown in an agent-based model that heterogeneity in *infectiousness* has a considerable impact on the feasibility of COVID-19 mitigation strategies (32). Our main finding is that another type of heterogeneity, namely the social kind, has a similarly profound effect on the effectiveness of contact tracing (Fig. 4): Structured social networks make mitigation by tracing much more cost-effective. Correspondingly, models which neglect social clustering are likely to underestimate the feasibility of contact tracing schemes.

We explore the effects of two central parameters, the *testing probability* and the *contact threshold* on the contact tracing scheme. The testing probability is influenced both by factors which are within our control, such as the overall availability of testing, and by factors which are essentially intrinsic to SARS-CoV-2, such as the rate at which symptoms develop. We find a non-trivial relation between testing probability and quarantine fraction, with a peak in quarantined individuals occurring at around 10% testing probability. The contact threshold, on the other hand, is a controllable parameter and essentially constitutes a design decision when e.g. developing contact tracing applications (20). We conclude that contact tracing remains effective, even if only relatively frequent contacts are quarantined.

## Data Availability

Datasets can be made available in GDPR-compliant form upon reasonable request.

## References

1. WO Kermack, AG McKendrick, GT Walker, A contribution to the mathematical theory of epidemics. Proc. Royal Soc. London. Ser. A, Containing Pap. a Math. Phys. Character 115, 700–721 (1927).

2. Norwegian Institute of Public Health, Coronavirus modelling at the NIPH (https://www.fhi.no/en/id/infectious-diseases/coronavirus/coronavirus-modelling-at-the-niph-fhi/) (2020) [Online; accessed 28-May-2020].

3. L Peng, W Yang, D Zhang, C Zhuge, L Hong, Epidemic analysis of covid-19 in china by dynamical modeling (2020).

4. N Ferguson, et al., Report 9: Impact of non-pharmaceutical interventions (npis) to reduce covid19 mortality and healthcare demand. Imp. Coll. COVID-19 Response Team (2020).

5. J Mossong, et al., Social contacts and mixing patterns relevant to the spread of infectious diseases. PLoS medicine 5 (2008).

6. P Klepac, S Kissler, J Gog, Contagion! the bbc four pandemic–the model behind the documentary. Epidemics 24, 49–59 (2018).

7. P Klepac, et al., Contacts in context: large-scale setting-specific social mixing matrices from the bbc pandemic project. medRxiv (2020).

8. L Pellis, S Cauchemez, NM Ferguson, C Fraser, Systematic selection between age and household structure for models aimed at emerging epidemic predictions. Nat. communications 11, 1–11 (2020).

9. K Prem, et al., The effect of control strategies to reduce social mixing on outcomes of the COVID-19 epidemic in Wuhan, China: a modelling study. The Lancet Public Heal. (2020).

10. MJ Keeling, et al., Predictions of covid-19 dynamics in the uk: short-term forecasting and analysis of potential exit strategies. medRxiv (2020).

11. MS Lau, et al., Spatial and temporal dynamics of superspreading events in the 2014–2015 west africa ebola epidemic. Proc. Natl. Acad. Sci. 114, 2337–2342 (2017).

12. MJ Keeling, TD Hollingsworth, JM Read, The efficacy of contact tracing for the containment of the 2019 novel coronavirus (covid-19). medRxiv (2020).

13. RM Anderson, H Heesterbeek, D Klinkenberg, TD Hollingsworth, How will country-based mitigation measures influence the course of the covid-19 epidemic? The Lancet 395, 931–934 (2020).

14. J Hellewell, et al., Feasibility of controlling covid-19 outbreaks by isolation of cases and contacts. The Lancet Glob. Heal. (2020).

15. AJ Kucharski, et al., Effectiveness of isolation, testing, contact tracing and physical distancing on reducing transmission of sars-cov-2 in different settings. medRxiv (2020).

16. L Ferretti, et al., Quantifying sars-cov-2 transmission suggests epidemic control with digital contact tracing. medRxiv (2020).

17. A Eilersen, K Sneppen, Estimating cost-benefit of quarantine length for covid-19 mitigation. medRxiv (2020).

18. M Gasparek, M Racko, M Dubovsky, A stochastic, individual-based model for the evaluation of the impact of non-pharmacological interventions on covid-19 transmission in slovakia. medRxiv (2020).

19. A Aleta, et al., Modeling the impact of social distancing, testing, contact tracing and household quarantine on second-wave scenarios of the covid-19 epidemic. medRxiv (2020).

20. Apple Inc., Building an App to Notify Users of COVID-19 Exposure (https://developer.apple.com/documentation/exposurenotification/building_an_app_to_notify_users_of_covid-19_exposure) (2020) [Online; accessed 31-May-2020].

21. S Maslov, K Sneppen, Specificity and stability in topology of protein networks. Science 296, 910–913 (2002).

22. A Stopczynski, et al., Measuring large-scale social networks with high resolution. PloS one 9, e95978 (2014).

23. A Mollgaard, et al., Measure of node similarity in multilayer networks. PloS one 11 (2016).

24. V Sekara, S Lehmann, The strength of friendship ties in proximity sensor data. PLOS ONE 9, 1–8 (2014).

25. World Health Organization, Q&A on coronaviruses (COVID-19) (https://www.who.int/emergencies/diseases/novel-coronavirus-2019/question-and-answers-hub/q-a-detail/q-a-coronaviruses) (2020) [Online; accessed 28-May-2020].

26. Centers for Disease Control and Prevention, How COVID-19 Spreads (https://www.cdc.gov/coronavirus/2019-ncov/prevent-getting-sick/how-covid-spreads.html) (2020) [Online; accessed 28-May-2020].

27. A Stopczynski, AS Pentland, S Lehmann, Physical proximity and spreading in dynamic social networks. arXiv preprint 1509.06530 (2015).

28. Our World In Data, European Centre for Disease Prevention and Control, covid-19-data (Deaths) (https://github.com/owid/covid-19-data/) (2020).

29. MGM Gomes, et al., Individual variation in susceptibility or exposure to sars-cov-2 lowers the herd immunity threshold. medRxiv (2020).

30. Y Zhang, Y Li, L Wang, M Li, X Zhou, Evaluating transmission heterogeneity and super-spreading event of covid-19 in a metropolis of china. Int. J. Environ. Res. Public Heal. 17, 3705 (2020).

31. BM Althouse, et al., Stochasticity and heterogeneity in the transmission dynamics of sars-cov-2 (2020).

32. K Sneppen, L Simonsen, Impact of superspreaders on dissemination and mitigation of covid-19. medRxiv (2020).

